# SARS-CoV-2 Seroprevalence in Relation to Timing of Symptoms

**DOI:** 10.1101/2020.08.02.20166876

**Authors:** Joseph E. Ebinger, Gregory J. Botwin, Christine M. Albert, Mona Alotaibi, Moshe Arditi, Anders H. Berg, Aleksandra Binek, Patrick Botting, Justyna Fert-Bober, Jane C. Figueiredo, Jonathan D. Grein, Wohaib Hasan, Mir Henglin, Shehnaz K. Hussain, Mohit Jain, Sandy Joung, Michael Karin, Elizabeth H. Kim, Dalin Li, Yunxian Liu, Eric Luong, Dermot P.B. McGovern, Akil Merchant, Noah Merin, Peggy B. Miles, Trevor-Trung Nguyen, Koen Raedschelders, Mohamad A. Rashid, Celine E. Riera, Richard V. Riggs, Sonia Sharma, Kimia Sobhani, Sarah Sternbach, Nancy Sun, Warren G. Tourtellotte, Jennifer E. Van Eyk, Jonathan G. Braun, Susan Cheng

**Affiliations:** Department of Cardiology, Cedars-Sinai Medical Center, Los Angeles, California, USA; Smidt Heart Institute, Cedars-Sinai Medical Center, Los Angeles, California, USA; F. Widjaja Foundation Inflammatory Bowel and Immunobiology Research Institute, Cedars-Sinai Medical Center, Los Angeles, California, USA; Division of Pulmonary and Critical Care Medicine, University of California, San Diego, San Diego, California, USA; Departments of Pediatrics, Division of Infectious Diseases and Immunology, and Infectious and Immunologic Diseases Research Center (IIDRC), Department of Biomedical Sciences, Cedars-Sinai Medical Center, Los Angeles, California, USA; Department of Pediatrics, David Geffen School of Medicine at UCLA, Los Angeles, California, USA; Department of Pathology and Laboratory Medicine, Cedars-Sinai Medical Center, Los Angeles, California, USA; Advanced Clinical Biosystems Institute, Department of Biomedical Sciences, Cedars-Sinai Medical Center, Los Angeles, California, USA; Cedars-Sinai Cancer and Department of Medicine, Cedars-Sinai Medical Center, Los Angeles, California, USA; Department of Medicine, Cedars-Sinai Medical Center, Los Angeles, California, USA; Department of Epidemiology, Cedars-Sinai Medical Center, Los Angeles, California, USA; Biobank & Translational Research Core Laboratory, Samuel Oschin Comprehensive Cancer Institute, Cedars-Sinai Medical Center, Los Angeles, California, USA; Department of Medicine, School of Medicine, University of California, San Diego, San Diego, CA; Department of Pharmacology, University of California, San Diego School of Medicine, San Diego, California, USA; Department of Internal Medicine, Division of Hematology Cedars-Sinai Medical Center, Los Angeles, California, USA; Employee Health Services, Department of Medicine, Cedars-Sinai Medical Center, Los Angeles, California, USA; Center for Neural Science and Medicine, Department of Biomedical Sciences, Board of Governors Regenerative Medicine Institute, Department of Neurology, Cedars-Sinai Medical Center, Los Angeles, California, USA; David Geffen School of Medicine, University of California, Los Angeles, Los Angeles, California, USA; Chief Medical Officer, Cedars-Sinai Medical Center, Los Angeles, California, USA; La Jolla Institute for Allergy and Immunology, La Jolla, California, USA; Barbra Streisand Women’s Heart Center, Cedars-Sinai Medical Center, Los Angeles, California, USA

## Abstract

Of individuals with SARS-CoV-2 IgG antibody testing performed, those who contemporaneously experienced a cluster of Covid-19 relevant symptoms in the 1-2 months preceding the antibody assay were more likely to test positive whereas those who experienced the symptom clustering in the prior 3-6 months were more likely to test negative. These findings suggest that antibodies likely wane over a period of months, particularly in relation to the timing of symptoms.

## Introduction

Serological studies that involve assaying circulating antibodies to SARS-CoV-2, the viral agent of COVID-19, represent a tractable approach to understanding patterns of exposure and potential immunity across populations at risk. Amidst legitimate concerns regarding the technical sensitivity and specificity of certain antibody testing kits, the application of validated assays has proved useful for estimating the probable prevalence of SARS-CoV-2 exposure in a given community at a particular point in time.^1-4^ Emerging evidence indicates that an antibody test result for a given individual also depends on when the test was performed in relation to onset of symptoms.^5^

## Methods

To clarify the relationship of antibody test results with the timing of symptoms, we analyzed data collected from a large cohort with SARS-CoV-2 serologic screening.^6^ Briefly, from May 11, 2020 to June 28, 2020 we enrolled N=6318 active employees working within the Cedars-Sinai Health System located in Los Angeles County, California. All participants were invited to receive SARS-CoV-2 IgG antibody testing (Abbott Architect^7^) in addition to completing an electronic survey of questions regarding Covid-19 related symptoms. Based on emerging data, the following 4 symptoms were considered most relevant to Covid-19: anosmia, fever, dry cough, and myalgias.^8,9^ All study protocols were approved by the Cedars Sinai institutional review board and all participants provided written informed consent. We used Pearson’s chi-square tests, one-way ANOVA, and multivariable regression analyses to examine potential differences in the timing of symptoms in relation to seroprevalence. All analyses were conducted using R v3.4.1 (R Foundation for Statistical Computing, Vienna, Austria).

## Results

A total of N=6062 individuals (age 41±12 years, 68% women, 19% Hispanic, 6% black) had complete data from both the serology testing and symptoms survey. Of those individuals who reported experiencing any symptoms potentially related to Covid-19 illness in the preceding 6 months (N=3688), a total of 176 (4.8%) tested positive for SARS-CoV-2 antibodies. Participants with antibodies to SARS-CoV-2, most frequently reported experiencing at least 1 of the 4 most relevant Covid-19 related symptoms during the 2 months preceding testing. By contrast, report of these same symptoms was most frequent during the 3 to 6 months preceding testing, among those without SARS-CoV-2 antibodies (**Figure 1A**).

**Figure 1A.**
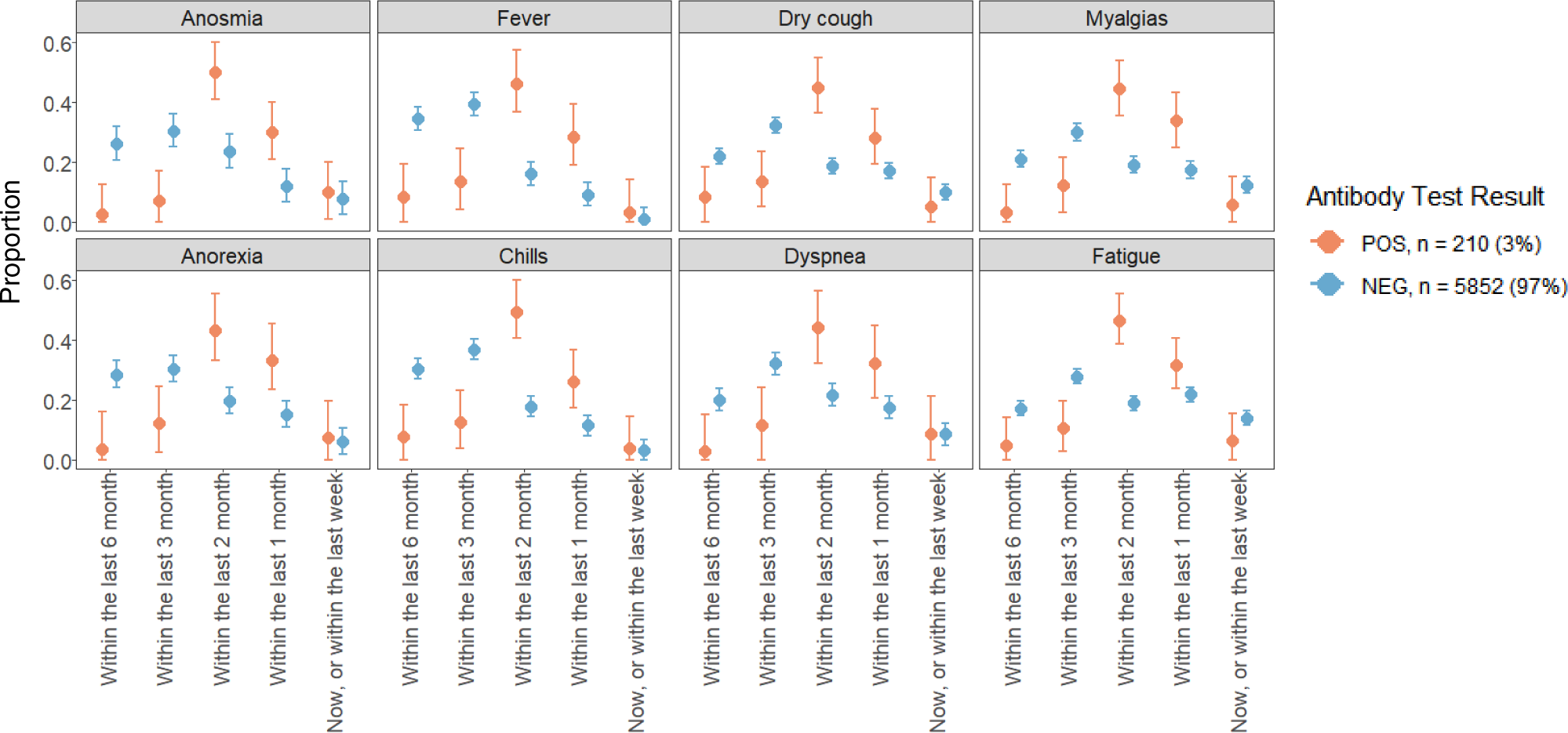
Timing of Symptoms Reported in Relation to SARS-CoV-2 IgG Antibody Test Result Status, in the Total Cohort.

Of the N=510 individuals who contemporaneously experienced at least 3 of the 4 most relevant symptoms, N=95 (19%) had SARS-CoV-2 antibodies (**Figure 1B**). Notably, among these participants, presence of this symptom cluster during the preceding 1 or 2 months was significantly associated with being seropositive (OR 14.6, 95% CI 8.4-26.6; P<0.001) in age- and sex-adjusted analyses. Conversely, having experienced this symptom cluster during the preceding 3 or 6 months was significantly associated with being seronegative (OR 19.4, 95% CI 10.7-37.7; P<0.001). Finally, N=158 participants reported receiving a medical diagnosis of Covid-19 prior to serology testing, of whom 95 (60.1%) did not have measurable SARS-CoV-2 antibodies: the seronegative group was more likely to experience their symptom cluster during the prior 3 to 6 months rather than within the prior 1 to 2 months leading up to antibody testing (OR 6.07, 95% CI 2.64-15.86; P<0.001).

**Figure 1B.**
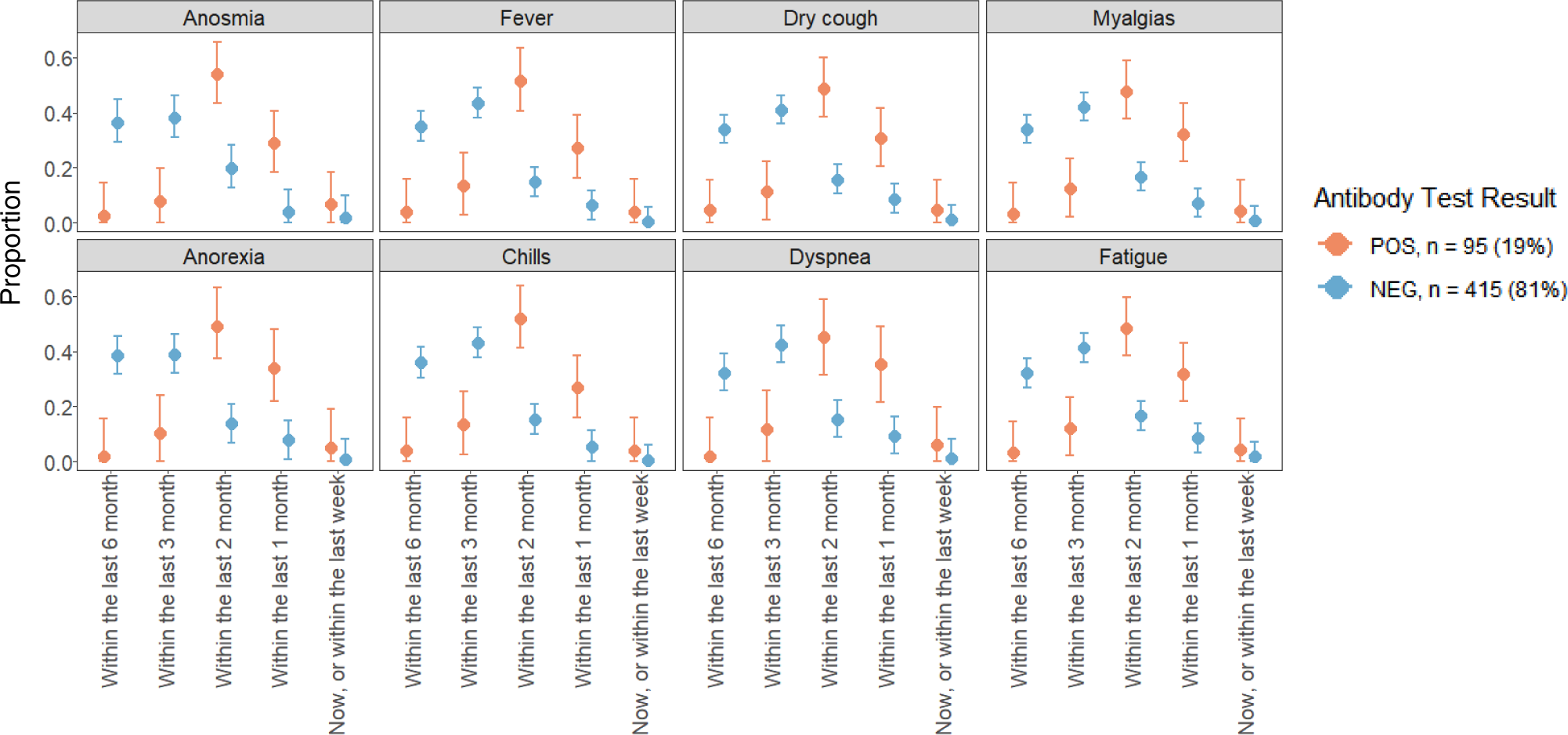
Timing of Symptoms Reported in Relation to SARS-CoV-2 IgG Antibody Test Result Status, Among Individuals Reporting Concurrent Clustering of Symptoms.

## Discussion

Across a large diverse cohort of individuals employed in a major metropolitan health system, a substantial proportion with detectable SARS-CoV-2 IgG antibodies experienced a contemporaneous cluster of COVID-19 relevant symptoms (including anosmia, fever, dry cough, and myalgias) at different timepoints leading up to the serology testing. Importantly, we found that symptoms were temporally associated with serologic status. Specifically, those who experienced Covid-19 symptoms within 2 months prior to testing were more likely to have SARS-CoV-2 antibodies, while those with symptoms in the 3-6 months prior to testing were more likely to be seronegative. Our findings are consistent with the known limited half-life of circulating IgG antibodies to other viral agents,^10^ in addition to emerging results from prior smaller studies that have directly observed SARS-CoV-2 IgG antibodies waning over a period of months.^11,12^ Our study of potential Covid-19 symptoms is limited by the similarity between symptoms relevant to Covid-19 and other viral pathogens, such as influenza, which was also present in the community 3-6 months prior to our study. We attempted to control for this limitation by focusing on symptom clustering that is more specific to Covid-19, including anosmia. Further investigations are needed to validate our results and also examine the extent to which antibody persistence or lack thereof may or may not represent time-dependent variation in relative immunity to Covid-19. Overall, our study findings offer important context for clinicians, scientists, and public health officials as part of ongoing efforts to deploy and interpreting results of antibody testing in populations at large.

## Data Availability

The data that support the findings of this study are available from Cedars-Sinai Medical Center, upon reasonable request. The data are not publicly available due to the contents including information that could compromise research participant privacy/consent.

## ACKNOWLEDGEMENTS

We are grateful to all the front-line healthcare workers in our healthcare system who continue to be dedicated to delivering the highest quality care for all patients.

## REFERENCES

1. Stringhini, S., et al. Seroprevalence of anti-SARS-CoV-2 IgG antibodies in Geneva, Switzerland (SEROCoV-POP): a population-based study. The Lancet.

2. Pollán, M., et al. Prevalence of SARS-CoV-2 in Spain (ENE-COVID): a nationwide, population-based seroepidemiological study. The Lancet.

3. Sood, N., et al. Seroprevalence of SARS-CoV-2–Specific Antibodies Among Adults in Los Angeles County, California, on April 10-11, 2020. JAMA 323, 2425–2427 (2020).

4. Xu, X., et al. Seroprevalence of immunoglobulin M and G antibodies against SARS-CoV-2 in China. Nature Medicine (2020).

5. Sethuraman, N., Jeremiah, S.S. & Ryo, A. Interpreting Diagnostic Tests for SARS-CoV-2. JAMA 323, 2249–2251 (2020).

6. Ebinger, J.E., et al. An Opportune and Relevant Design for Studying the Health Trajectories of Healthcare Workers. medRxiv, 2020.2006.2030.20140046 (2020).

7. Bryan, A., et al. Performance Characteristics of the Abbott Architect SARS-CoV-2 IgG Assay and Seroprevalence in Boise, Idaho. J Clin Microbiol (2020).

8. Wise, J. Covid-19: Study reveals six clusters of symptoms that could be used as a clinical prediction tool. BMJ 370, m2911 (2020).

9. Steensels, D., et al. Hospital-Wide SARS-CoV-2 Antibody Screening in 3056 Staff in a Tertiary Center in Belgium. JAMA (2020).

10. Krammer, F. The human antibody response to influenza A virus infection and vaccination. Nature Reviews Immunology 19, 383–397 (2019).

11. Long, Q.-X., et al. Clinical and immunological assessment of asymptomatic SARS-CoV-2 infections. Nature Medicine (2020).

12. Ibarrondo, F.J., et al. Rapid Decay of Anti–SARS-CoV-2 Antibodies in Persons with Mild Covid-19. New England Journal of Medicine (2020).

